# Risk Factors of the Severity of COVID-19: a Meta-Analysis

**DOI:** 10.1101/2020.04.30.20086744

**Authors:** Abdur Rahman, Nusrat Jahan Sathi

**Author notes:** CORRESPONDING AUTHOR: Abdur Rahman, Statistics Discipline, Khulna University, Khulna-9208, Bangladesh.

## Abstract

**BACKGROUND:** Although the infection rate of COVID-19 is very high, all the patients getting infected don’t always die or go through brutal states. This indicates there may be some factors that possibly boost the severity of COVID-19.

**OBJECTIVE:** We intend to identify some probable risk factors that are responsible for the severity of COVID-19 using a meta-analysis.

**METHODS:** The literature exploration lasted up to 18 April 2020 and through PubMed, Google Scholar, EMBASE, and Cochrane Library we have identified 10 pertinent publications. To paraphrase the outcomes of autonomous researches, we have performed a random-effect meta-analysis.

**RESULTS:** A total of 2272 patients’ information was extracted from the selected literature. We have found sex (male) (Risk ratio [RR] =1.29 [1.07; 1.54]), hypertension (RR=1.79 [1.57; 2.04]), diabetes (RR=1.57 [1.25; 1.98]), fatigue or myalgia (RR=1.17 [1.02; 1.35]), and smoking history (RR=1.71 [1.25; 2.35]) are potential risk factors for the severity of COVID-19. We found fever (RR=1.21 [0.66; 2.22]), cough (1.13 [0.98; 1.30]), and diarrhea (RR=1.14 [0.93; 1.40]) as insignificant risk factors for COVID-19 severity.

**CONCLUSION:** The findings of this research may be beneficial to identify patients with higher risks to provide additional medical attention from the very beginning of the treatment.

## INTRODUCTION

The coronavirus disease 2019 (COVID-19) originated from Wuhan (Hubei state, China), carrying similar DNA structure to SARS (Severe Acute Respiratory Syndrome) and MERS (Middle East Respiratory Syndrome) has spread throughout the world and creating massive panic to the human life^1,2^. The disease has the worst feature to transmit from person to person^3^, considering this feature and its lofty infection rate on January 30, 2020, the World Health Organization (WHO) declared COVID-19 as a global emergency.

To date, it has infected more than 3 million people and over 2 lakh have died. The outbreak has hit the USA, Italy, and Spain very badly. This three-country together holds over 1.3 million total identified cases and over 50% of the total death because of COVID-19^4^. As no proven treatment/medicine or vaccine is available to date^5^ the harm of COVID-19 has already overtaken SARS and MARS^6^.

Although the infection rate is very high, all the patients getting infected by this disease don’t always die. The global recovery rate is about 28.6% and the death rate is about 7% until April 28, 2020^4^. This information suggests that there may be some factors that influence the risk of death or critical medical states of the patients. That’s why it is important to identify and estimate such risk factors to predict the severe complication of the patients for avoiding or to minimize the severity^7^.

Researchers are trying to identify risk factors that deteriorate the health state of the COVID-19 patients mostly by using meta-analysis and systematic review. Some earlier investigations reported males are more likely to die or to go through the critical states of COVID-19^8,9^. There is an ongoing debate on whether smoking is a risk factor for COVID-19 severity. Although some regard it as a risk factor^9,27^, others found no significant alliance between smoking and the severity of COVID-19^8,10^. Other clinical traits termed as risk factors in the publications are hypertension^11,12^, diabetes^12,13^, and fever^8^.

In this study, we endeavor to identify some demographic and clinical characteristics which can be appraised as risk factors for the severity of COVID-19 by summarizing findings of the published literature.

## METHODS

### LITERATURE SEARCH

The literature search lasted from April 2, 2020, to April 18, 2020. Both of the authors searched through PubMed, Google Scholar, EMBASE, and Cochrane Library using keywords: “COVID-19”, “Novel Coronavirus”, “COVID-19 characteristics”, “COVID-19 patient”, and “China coronavirus”. We initially identified all the studies conformed to the keywords without any further investigations.

### INCLUSION CRITERIA

Based on the following criteria, we have included literature in the current study: a) bivariate data available for the severity (death/ICU (Intensive Care Unit)/severe state/others) of COVID-19 patients, b) multiple factors available for the severity of the disease, c) full-text access to the article, d) information presented in English language, e) peer-reviewed accepted/published articles, and f) literature published in and after December 2019 (after the first patient identified). We illustrate the systematic selection procedure of literature in Figure 1.

**Figure 1:**
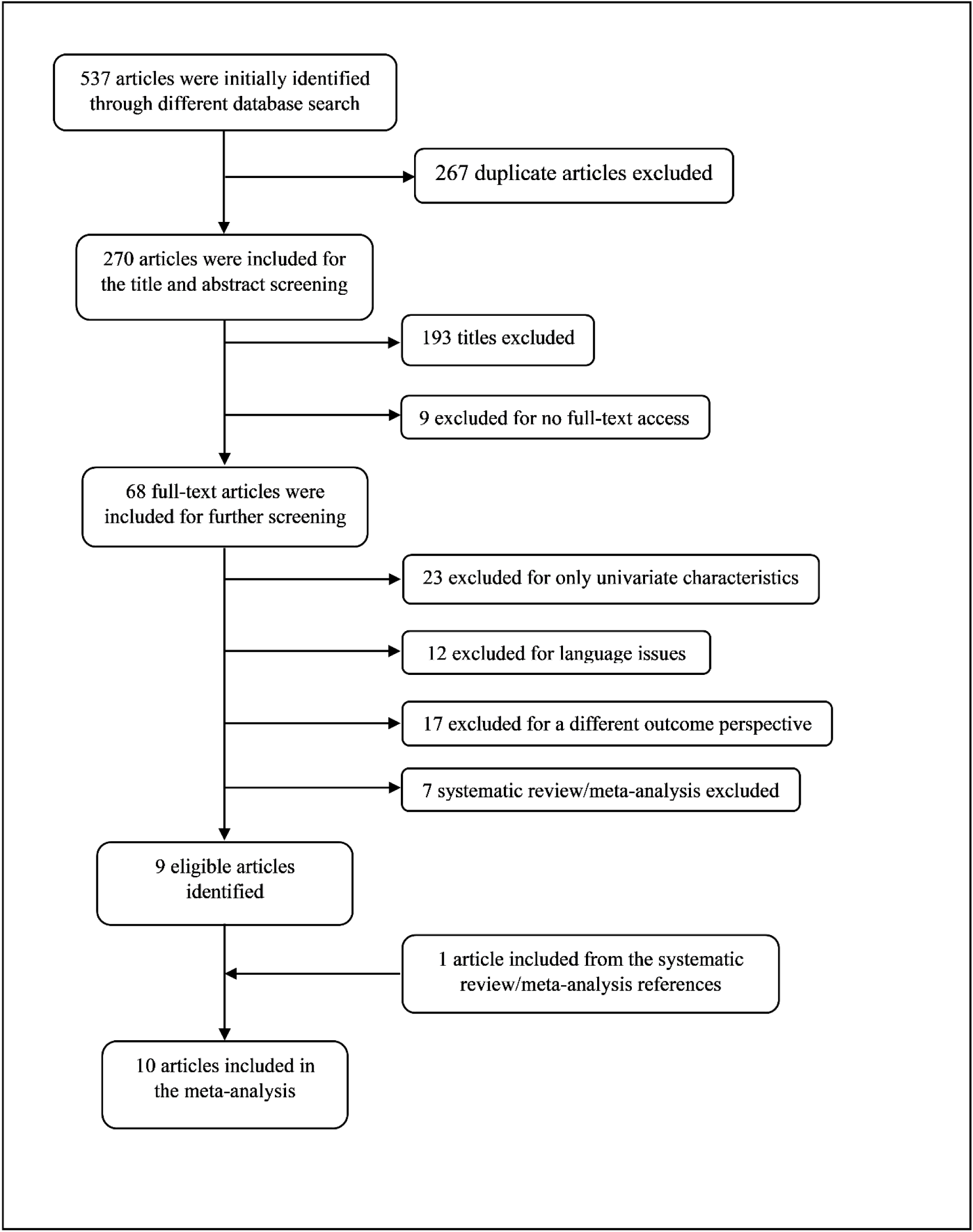
Flow chart showing the study selection procedure

### OUTCOMES AND COVARIATES

The authors extracted the number of patients that have undergone the severity of COVID-19 and are stable/recovered under various factors like sex, smoking history, fatigue or myalgia, cough, fever, diarrhea, hypertension, and diabetes. The term severity in this study represents death/ICU/severe state or any other critical medical state. We also extracted the authors’ names, publication year, study design, sample size, country, and other related information from the finally selected studies.

### STATISTICAL ANALYSIS

Considering the primary goal of the study we performed random effect meta-analysis where the effect sizes were computed from the raw information dragged from individual studies. We considered the effect size for the study to be a Risk Ratio [RR] and reported with a 95% confidence interval. I^2^ statistic reported in the forest plot reflects relative between studies heterogeneity and P value (two tail with level 0.05) determines the significance of the heterogeneity test. Microsoft Excel 2013 was used to create metadata, get their univariate characteristics, and export to other software for further analysis. Random effect meta-analysis and relevant forest plots were generated using statistical software R version 3.5.1.

## RESULTS

We have extracted 2272 patients’ information from 10 autonomous studies^14–23^ and incorporated them into this research. The highest sample size for the isolated study was 1099, and 34 was the smallest sample size, other study characteristics are summarized in Table 1.

**Table 1:** Characteristics of the selected literatures

| Author | Population with Placing | Size | Study Design | Time Period | Outcome | Factors included in the study |
| --- | --- | --- | --- | --- | --- | --- |
| Chen et al<sup>14</sup> | Covid-19 positive patient from Tongji Hospital in Wuhan, China | 274 | Retrospective study | Jan 13, 2020 to Feb 12, 2020 | Death | Sex, smoke, diabetes, hypertension, fever, cough, fatigue or myalgia, and diarrhea |
| Deng et al<sup>15</sup> | Patients from two tertiary hospitals in Wuhan, China | 225 | Retrospective study | Jan 1, 2020 to Feb 21, 2020 | Death | Sex, diabetes, hypertension, fever, cough, fatigue, and diarrhea |
| Guan et al<sup>16</sup> | Confirmed patients from 552 hospitals in 30 provinces, China | 1099 | Retrospective study | Dec 11, 2019 to Jan 29, 2020 | Severe | Sex, smoking history, diabetes, hypertension, fever, cough, fatigue or myalgia, and diarrhea |
| Huang et al<sup>17</sup> | All Covid-19 patients from a designed hospital in Wuhan, China | 41 | Retrospective study | Dec 16, 2019 to Jan 2, 2020 | ICU | Sex, smoking history, diabetes, hypertension, fever, cough, fatigue or myalgia, and diarrhea |
| Lei et al<sup>18</sup> | Clinical data from Renmin, Zhongnan, Tongji and Central Hospital, China | 34 | Retrospective study | Jan 1, 2020 to Feb 5, 2020 | ICU | Sex, diabetes, hypertension, fever, cough, fatigue or myalgia, and diarrhea |
| Liu et al<sup>19</sup> | Patients from three tertiary hospitals in Wuhan, China | 78 | Retrospective study | Dec 30, 2019 to Jan 15, 2020 | Progression | Sex, smoking history, hypertension, diabetes, and cough |

Table 1: Characteristics of the selected literatures (Continue)
| Author | Population with Placing | Size | Study Design | Time Period | Outcome | Factors included in the study |
| --- | --- | --- | --- | --- | --- | --- |
| Wang et al<sup>20</sup> | Consecutive hospitalized patient at Zhongnan Hospital in Wuhan, China | 138 | Retrospective study | Jan 1, 2020 to Jan 28, 2020 | ICU | Sex, diabetes, hypertension, fever, cough, fatigue or myalgia, and diarrhea |
| Yang et al<sup>21</sup> | Adult patients in Wuhan Jin Yin-tan hospital, China | 52 | Retrospective study | Dec 2019 to Jan 26, 2020 | Non-survivor | Sex, smoking history, diabetes, fatigue or myalgia, hypertension, and cough |
| Zhang et al<sup>22</sup> | All hospitalized patients from Wuhan hospital, China | 140 | Retrospective study | Jan 5, 2020 to Jan 24, 2020 | Severe | Sex, smoking history, diabetes, hypertension, fever, cough, fatigue or myalgia, and diarrhea |
| Zhou et al<sup>23</sup> | Adult ($\geq 18$ years) patients from Jinyintan Hospital and Wuhan Pulmonary Hospital, China | 191 | Retrospective multi-center cohort study | Dec 29, 2019 to Jan 31, 2020 | Non-survivor | Sex, smoking history, diabetes, hypertension, fever, cough, fatigue or myalgia, and diarrhea |

Table 2 demonstrates that the bulk of the COVID-19 patients included in the study through a methodical scheme are male (60%). The clinical characteristics of the patients comprise diabetes (11.2%), hypertension (21.3%), fever (90.2%), cough (65%), fatigue or myalgia (41.0%), and diarrhea (9.2%). Only 11.3% of the patients smoke cigarettes (current or past smoker).

**Table 2:** Baseline characteristics of COVID-19 patients for different studies

| Characteristics | Chen et al<sup>14</sup> | Deng et al<sup>15</sup> | Guan et al<sup>16</sup> | Huang et al<sup>17</sup> | Lei et al<sup>18</sup> | Liu et al<sup>19</sup> | Wang et al<sup>20</sup> | Yang et al<sup>21</sup> | Zhang et al<sup>22</sup> | Zhou et al<sup>23</sup> | Overall |
| --- | --- | --- | --- | --- | --- | --- | --- | --- | --- | --- | --- |
| Male | 171<br>(62.4) | 124<br>(55.1) | 637<br>(58.1) | 30<br>(73.2) | 14<br>(41.2) | 39<br>(50.0) | 75<br>(54.3) | 35<br>(67.3) | 71<br>(50.7) | 119<br>(62.6) | 1315<br>(60.0) |
| Smoking history | 19<br>(6.9) | - | 158<br>(14.6) | 3<br>(7.3) | - | 5<br>(6.4) | - | - | 9<br>(6.4) | 11<br>(5.8) | 205<br>(11.3) |
| Diabetes | 47<br>(17.2) | 26<br>(11.6) | 81<br>(7.4) | 8<br>(19.5) | 8<br>(23.5) | 5<br>(6.4) | 14<br>(10.1) | 9<br>(17.3) | 17<br>(12.1) | 36<br>(22.6) | 251<br>(11.2) |
| Hypertension | 93<br>(33.9) | 58<br>(25.8) | 165<br>(15.0) | 6<br>(14.6) | 13<br>(38.2) | 8<br>(10.3) | 43<br>(31.2) | - | 42<br>(30.0) | 58<br>(30.4) | 468<br>(21.3) |
| Fever | 249<br>(90.9) | 189<br>(84.0) | 975<br>(88.7) | 40<br>(97.6) | 31<br>(91.2) | - | 136<br>(98.6) | 51<br>(98.1) | 110<br>(91.7) | 180<br>(94.2) | 1961<br>(90.2) |
| Cough | 185<br>(67.5) | 85<br>(37.8) | 745<br>(67.8) | 31<br>(75.6) | 18<br>(52.9) | 34<br>(43.6) | 82<br>(59.4) | 40<br>(76.9) | 90<br>(75.0) | 151<br>(79.1) | 1461<br>(65.0) |
| Fatigue or myalgia | 137<br>(50.0) | 57<br>(25.3) | 419<br>(38.1) | 18<br>(43.9) | 25<br>(73.5) | - | 96<br>(69.6) | 6<br>(11.5) | 90<br>(75.0) | 44<br>(23.0) | 892<br>(41.0) |
| Diarrhea | 77<br>(28.1) | 33<br>(14.7) | 42<br>(3.8) | 1<br>(2.6) | 2<br>(5.9) | - | 14<br>(10.1) | - | 18<br>(12.9) | 9<br>(4.7) | 196<br>(9.2) |

Note: Data are presented as n, and (n/N \*100%); where n is the number of patients with certain characteristics and N is the total number of available patients observed in the study under that certain characteristics.

Forest plot in (Figure 2) illustrates that only six studies were entailed for the meta-analysis for characteristic smoking history where all ten studies availed information about the sex of the patients. From the random-effect meta-analysis, we see that patients with smoking history have a higher risk to experience a severe state of COVID-19 (RR =1.71; 95% CI, 1.25 to 2.35) or 71% higher risk pertained to a non-smoker patient. The pooled risk ratio for the male suggests higher risk compared to female patients (RR=1.29; 95% CI, 1.07 to 1.54). In both cases between-study heterogeneity is low and the test of heterogeneity is insignificant (I^2^=38%, P=0.15; and I^2^=33%, P=0.15, respectively).

**Figure 2:**
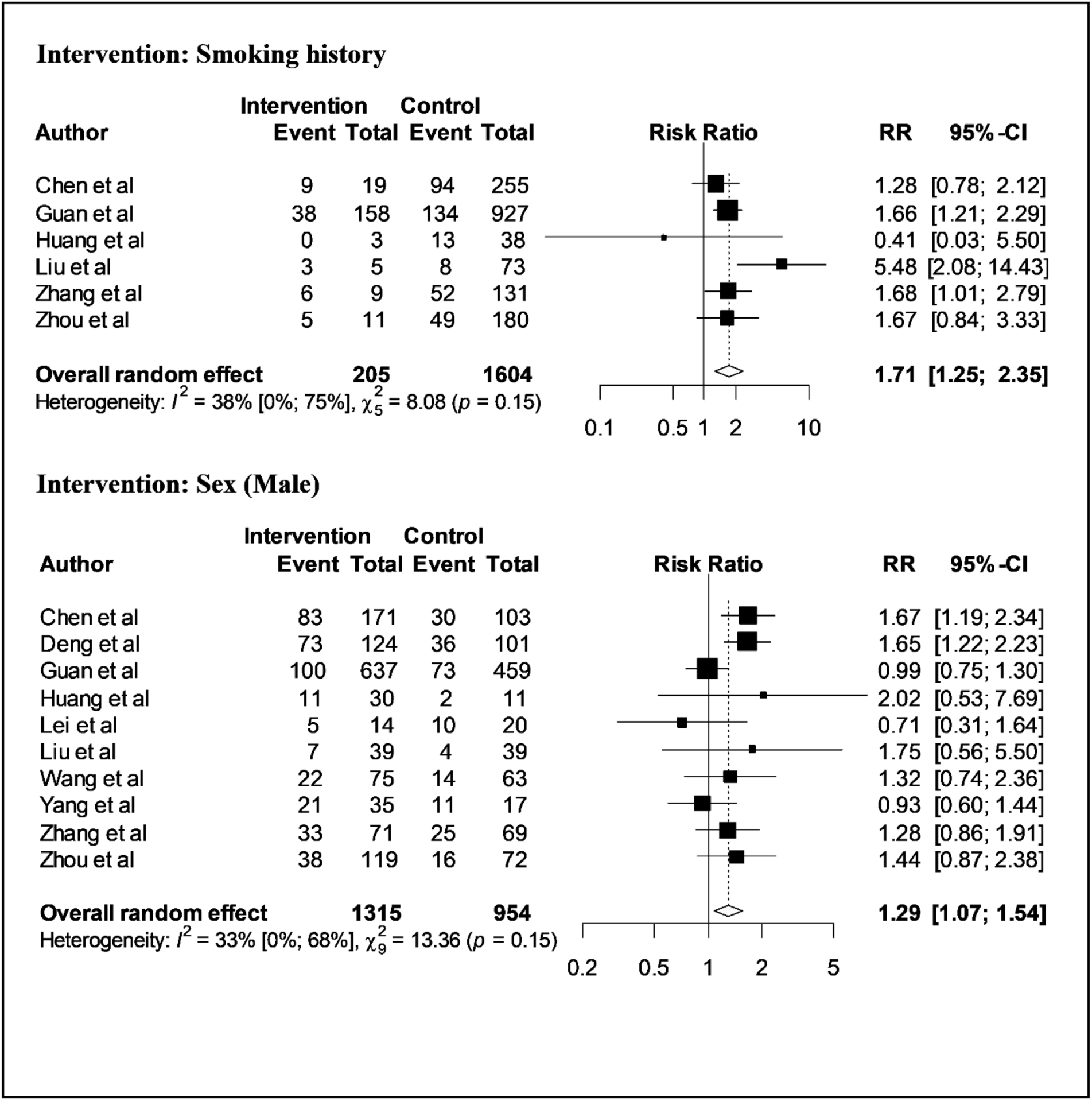
Forest plot for factors smoking history and sex (male) illustrating the distribution of the risk ratio of COVID-19 severity

Figure 3 displays individual and pooled risk ratios for patients with diabetes and hypertension. Although six isolated studies data implies that diabetes is an insignificant risk factor but the overall effect is significant and indicates a higher risk to undergo brutal state for a patient with diabetes (RR =1.57; 95% CI, 1.25 to 1.98). The heterogeneity is moderate (I^2^=54%) and the test for heterogeneity is significant (*χ*^2^=19.59, P=0.02). Also, patients with hypertension have a higher risk (RR=1.79; 95% CI, 1.57 to 2.04) compared to patient those are not suffering from hypertension with insignificant heterogeneity, I^2^=0% (P=0.50).

**Figure 3:**
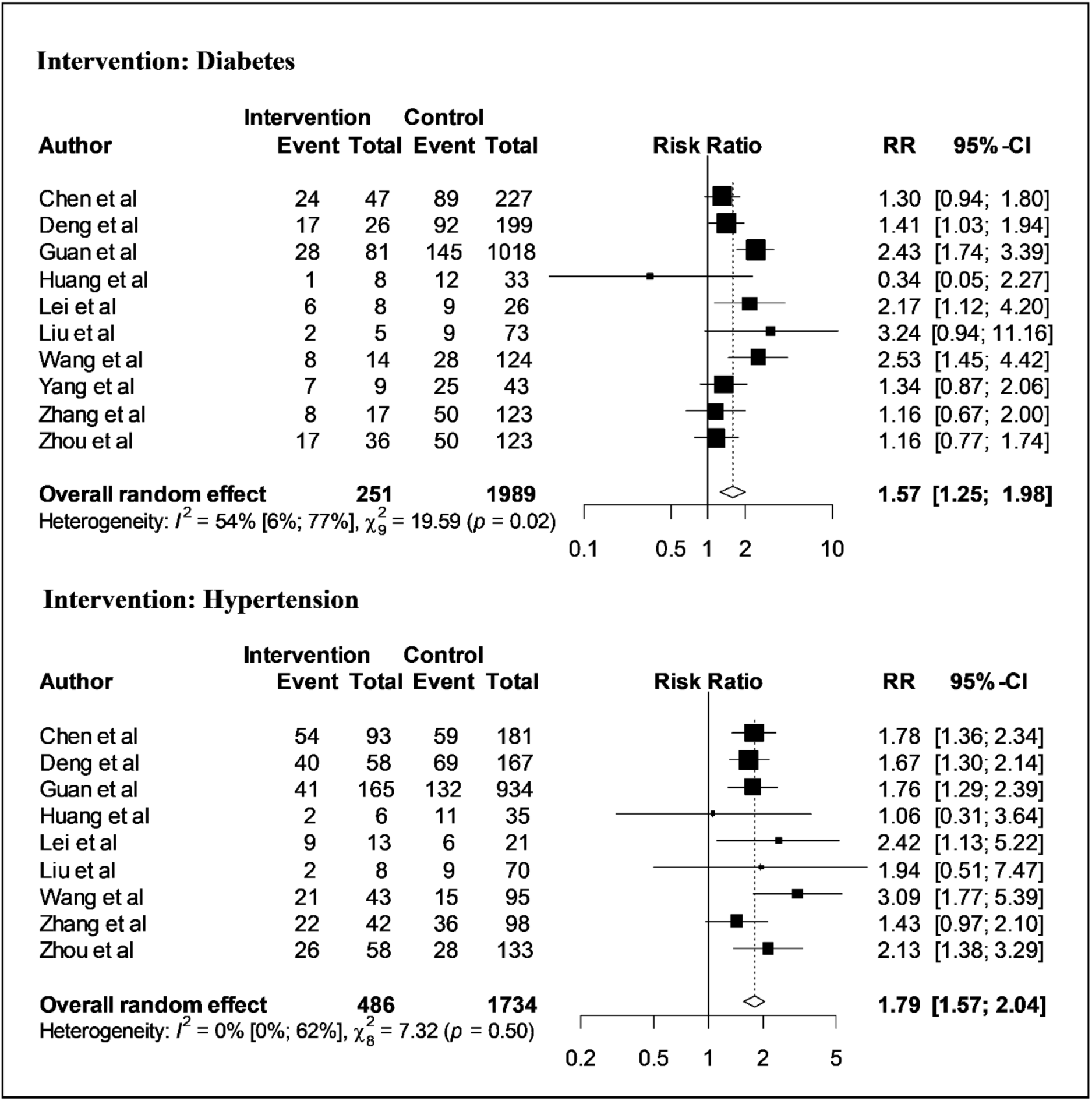
Forest plot for factors diabetes and hypertension illustrating the distribution of the risk ratio of COVID-19 severity

Respectively eight and nine studies were involved in the analysis for factors diarrhea and fatigue or myalgia of patients (Figure 4). In both cases the heterogeneity is very low and is insignificant (I^2^=8%, P=0.37) and (I^2^=0%, P=0.86), respectively). The pooled random effect infers diarrhea have an insignificant impact on the severity of COVID-19 (RR=1.14; 95% CI, 0.93 to 1.40). But the overall RR=1.17 (95% CI, 1.02 to 1.35) suggests that patients with fatigue or myalgia have a 17% higher risk to go through a severe state of COVID-19.

**Figure 4:**
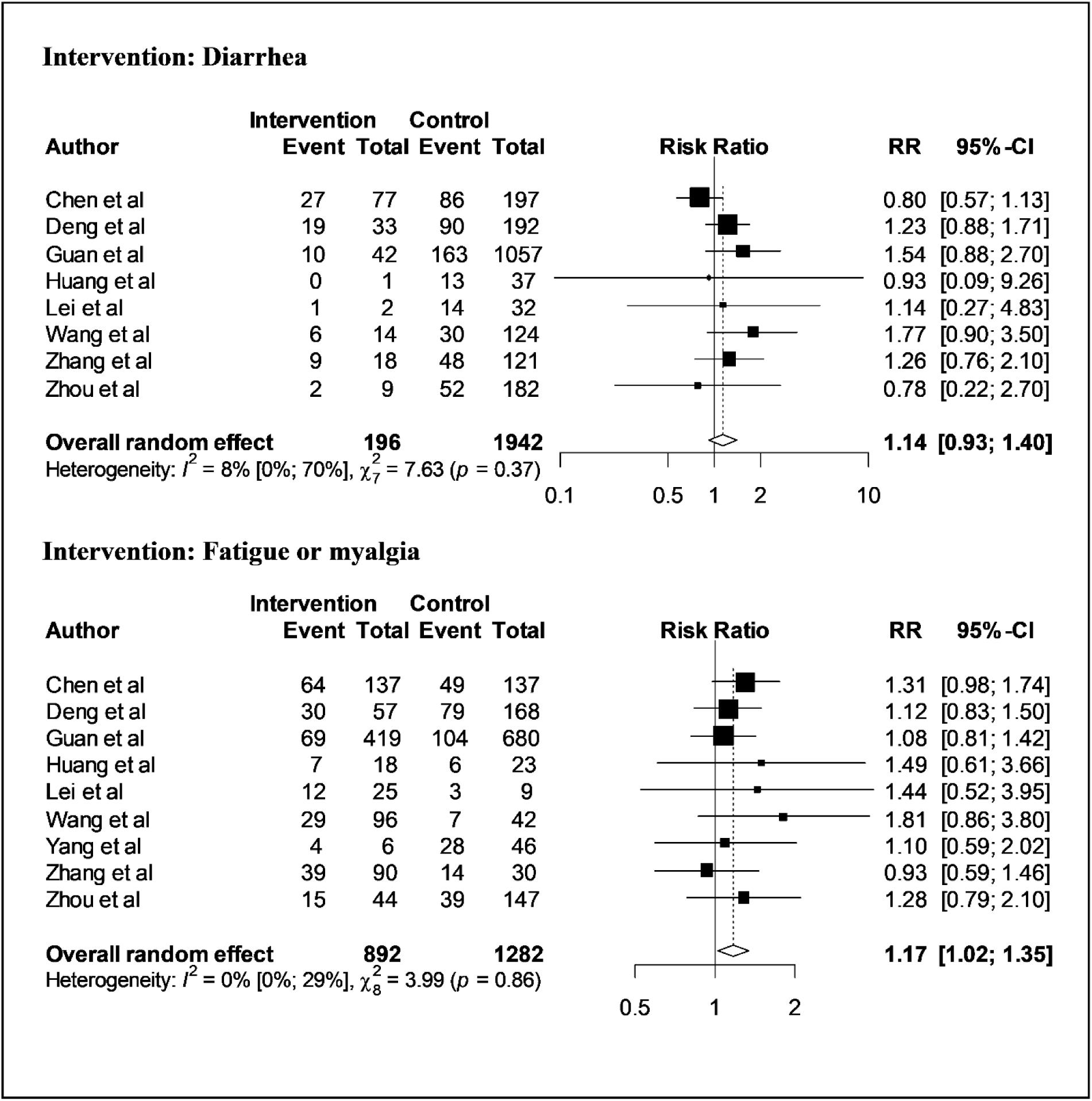
Forest plot for factors diarrhea and fatigue or myalgia illustrating the distribution of the risk ratio of COVID-19 severity

From (Figure 5), we glimpse that cough and fever have no considerable relation with the severity of COVID-19. The pooled effect reflects patient with cough symptom has RR=1.13 (95% CI, 0.98 to 1.30) and patient with fever has RR=1.21 (95% CI, 0.66 to 2.22). Although for cough symptom the between-study heterogeneity is nil and insignificant (I^2^=0%, P=0.48), it is very big and also significant for fever (I^2^=85%, P<0.01).

**Figure 5:**
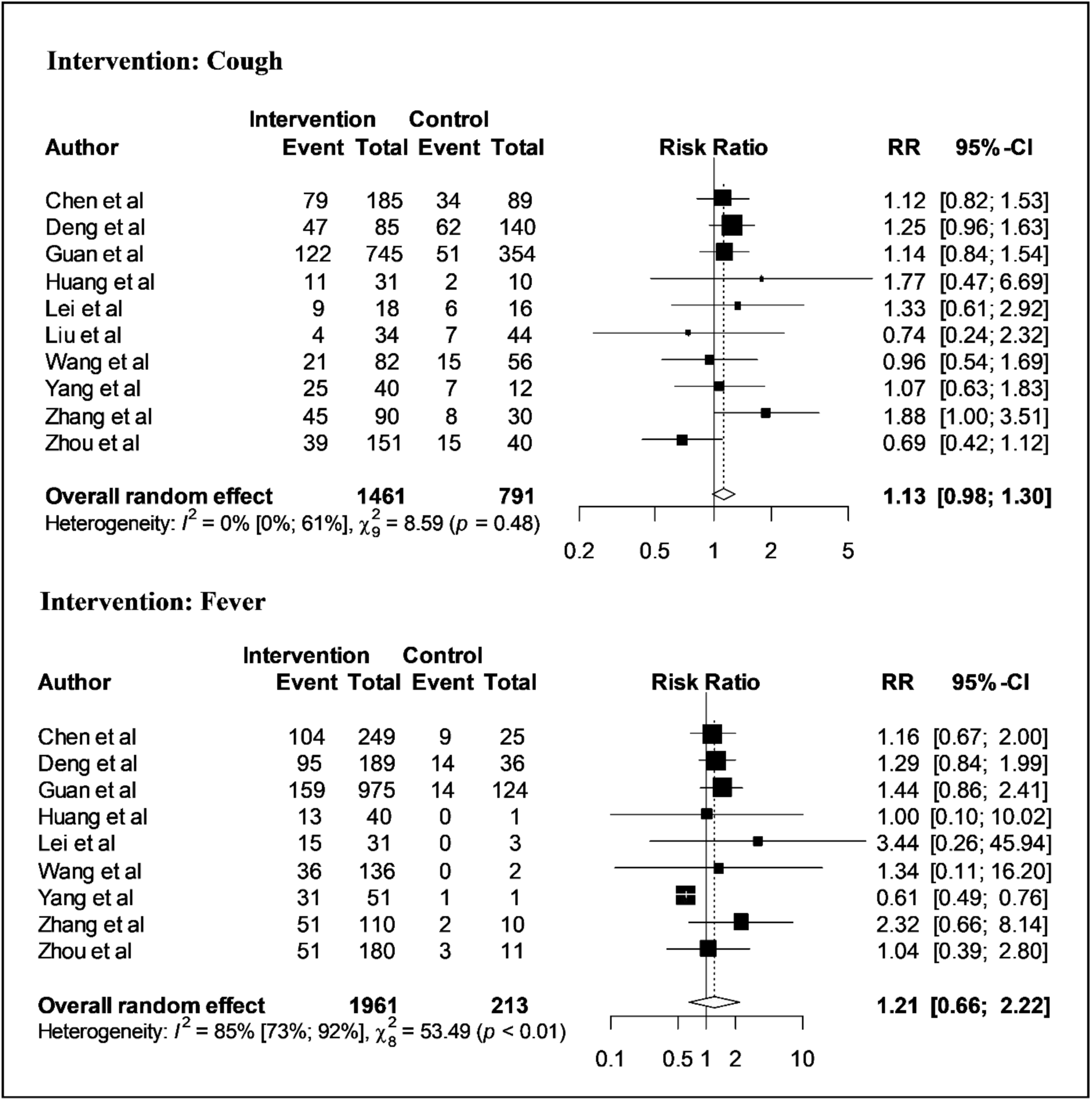
Forest plot for factors cough and fever illustrating the distribution of the risk ratio of COVID-19 severity

## DISCUSSION

This study through an organized way incorporated 10 publications and performed a random-effect meta-analysis to identify some risk factors that are probably accountable for the severity of COVID-19. Although none of the articles incorporated in the current research reports any effect size, we computed the effect size (risk ratio) from the published data. The pooled effect size with confidence interval deemed to avouch a factor as a significant risk factor.

We found male patients have a reasonably higher risk to undergo the severity of COVID-19 compared to female patients echoing with some previous studies^8,9^. An explanation of this can be, the male has weaker immunity because of genetic and hormonal factors and has shown higher mortality in several infectious diseases^24,25^.

Although some aforementioned studies^8,10^ found smoking as an insignificant factor to worsen the state of COVID-19 patients, we have found smoking as a significant factor. Smoking is associated with lower or impoverished immunity of smokers^26^, which can explain why smokers might have elevated risks. Some previous studies^9,27^ supports our finding, they documented smoking as a significant risk factor, and smokers have a higher risk to undergo a severe state of COVID-19. In a previous study^28^, smoking was also reported as a risk factor for the critical illness of MERS (Middle East Respiratory Syndrome) too.

We incorporated some clinical traits of the patients like cough, fever, fatigue or myalgia, diarrhea, diabetes, and hypertension in this study. Among those, we have found fatigue or myalgia, hypertension, and diabetes are significant risk factors that might lead to a critical state of COVID-19 patients. Some previous studies^8,12,13^ also reported both diabetes and hypertension are significant risk factors of COVID-19 progression. These two comorbidities negatively affected MERS-CoV patients’ too^29^.

From the random-effect meta-analysis, we conclude fever, cough, and diarrhea are insignificant risk factors for the severity of COVID-19. But a previous meta-analysis^8^ published fever as a significant risk factor for the severity of COVID-19.

## CONCLUSIONS

In conclusion, we consider sex (male), smoking history, diabetes, hypertension, and fatigue or myalgia are significant risk factors for the severity of COVID-19. It may require additional medical attention for patients with higher risk from the very beginning of the treatment. We also hope the findings of this study may assist experts in vaccine development programs.

## Data Availability

The studies are available online and full-text accessible.

## FUNDING

This research did not receive any specific grant from funding agencies in the public, commercial, or not-for-profit sectors.

## CONFLICT OF INTEREST

None declared.

## Notes

### Competing Interest Statement

The authors have declared no competing interest.

### Funding Statement

No funding

## References

1. Chen N, Zhou M, Dong X, Qu J, Gong F, Han Y, Qiu Y, Wang J, Liu Y, Wei Y, Yu T. Epidemiological and clinical characteristics of 99 cases of 2019 novel coronavirus pneumonia in Wuhan, China: a descriptive study. The Lancet. 2020 Feb 15;395(10223):507–13. (https://doi.org/10.1016/S0140-6736(20)30211-7)

2. Surveillances V. The epidemiological characteristics of an outbreak of 2019 novel coronavirus diseases (COVID-19)—China, 2020. China CDC Weekly. 2020;2(8):113–22.

3. Grabinskinski LE, Menachery VD. Return of the Coronavirus: 2019-nCoV. Viruses. 2020 Feb;12(2):135.

4. Worldometers. info, Dover, Delaware, U.S.A [Cited 28 April 2020] (http://www.worldometers.info/coronavirus/#countries)

5. Heymann DL, Shindo N. COVID-19: what is next for public health?. The Lancet. 2020 Feb 22;395(10224):542–5. (https://doi.org/10.1016/S0140-6736(20)30374-3)

6. Lu H, Stratton CW, Tang YW. Outbreak of Pneumonia of Unknown Etiology in Wuhan China: the Mystery and the Miracle. J Med Virol 2020. (https://doi.org/10.1002/jmv.25678)

7. Zumla A, Hui DS, Perlman S. Middle East respiratory syndrome. The Lancet. 2015 Sep 5;386(9997):995–1007.

8. Xu L, Chen G. Risk factors for severe corona virus disease 2019 (COVID-19) patients: a systematic review and meta analysis. medRxiv. 2020 Jan 1.

9. Zhao X, Zhang B, Li P, Ma C, Gu J, Hou P, Guo Z, Wu H, Bai Y. Incidence, clinical characteristics and prognostic factor of patients with COVID-19: a systematic review and metaanalysis. medRxiv. 2020 Jan 1.

10. Lippi G, Henry BM. Active smoking is not associated with severity of coronavirus disease 2019 (COVID-19). European journal of internal medicine. 2020 Mar 16.

11. Yang, J., Zheng, Y., Gou, X., Pu, K., Chen, Z., Guo, Q., Ji, R., Wang, H., Wang, Y. and Zhou, Y., 2020. Prevalence of comorbidities in the novel Wuhan coronavirus (COVID-19) infection: a systematic review and meta-analysis. International Journal of Infectious Diseases.

12. Li B, Yang J, Zhao F, Zhi L, Wang X, Liu L, Bi Z, Zhao Y. Prevalence and impact of cardiovascular metabolic diseases on COVID-19 in China. Clinical Research in Cardiology. 2020 Mar 11: 1–8.

13. Guo W, Li M, Dong Y, Zhou H, Zhang Z, Tian C, Qin R, Wang H, Shen Y, Du K, Zhao L. Diabetes is a risk factor for the progression and prognosis of COVID-19. Diabetes/Metabolism Research and Reviews. 2020 Mar 31.

14. Chen T, Wu D, Chen H, Yan W, Yang D, Chen G, Ma K, Xu D, Yu H, Wang H, Wang T. Clinical characteristics of 113 deceased patients with coronavirus disease 2019: retrospective study. Bmj. 2020 Mar 26;368.

15. Deng Y, Liu W, Liu K, Fang YY, Shang J, Wang K, Leng F, Wei S, Chen L, Liu HG. Clinical characteristics of fatal and recovered cases of coronavirus disease 2019 (COVID-19) in Wuhan, China: a retrospective study. Chinese Medical Journal. 2020 Mar 27.

16. Guan WJ, Ni ZY, Hu Y, Liang WH, Ou CQ, He JX, Liu L, Shan H, Lei CL, Hui DS, Du B. Clinical characteristics of coronavirus disease 2019 in China. New England Journal of Medicine. 2020 Feb 28.

17. Huang C, Wang Y, Li X, Ren L, Zhao J, Hu Y, Zhang L, Fan G, Xu J, Gu X, Cheng Z. Clinical features of patients infected with 2019 novel coronavirus in Wuhan, China. The Lancet. 2020 Feb 15;395(10223):497–506.

18. Lei S, Jiang F, Su W, Chen C, Chen J, Mei W, Zhan LY, Jia Y, Zhang L, Liu D, Xia ZY. Clinical characteristics and outcomes of patients undergoing surgeries during the incubation period of COVID-19 infection. EClinicalMedicine. 2020 Apr 5: 100331.

19. Liu W, Tao ZW, Wang L, Yuan ML, Liu K, Zhou L, Wei S, Deng Y, Liu J, Liu HG, Ming Y. Analysis of factors associated with disease outcomes in hospitalized patients with 2019 novel coronavirus disease. Chinese medical journal. 2020 Feb 28.

20. Wang D, Hu B, Hu C, Zhu F, Liu X, Zhang J, Wang B, Xiang H, Cheng Z, Xiong Y, Zhao Y. Clinical characteristics of 138 hospitalized patients with 2019 novel coronavirus–infected pneumonia in Wuhan, China. Jama. 2020 Mar 17;323(11):1061–9.

21. Yang X, Yu Y, Xu J, Shu H, Liu H, Wu Y, Zhang L, Yu Z, Fang M, Yu T, Wang Y. Clinical course and outcomes of critically ill patients with SARS-CoV-2 pneumonia in Wuhan, China: a single-centered, retrospective, observational study. The Lancet Respiratory Medicine. 2020 Feb 24.

22. Zhang JJ, Dong X, Cao YY, Yuan YD, Yang YB, Yan YQ, Akdis CA, Gao YD. Clinical characteristics of 140 patients infected with SARS-CoV-2 in Wuhan, China. Allergy. 2020 Feb

23. Zhou F, Yu T, Du R, Fan G, Liu Y, Liu Z, Xiang J, Wang Y, Song B, Gu X, Guan L. Clinical course and risk factors for mortality of adult inpatients with COVID-19 in Wuhan, China: a retrospective cohort study. The Lancet. 2020 Mar 11.

24. Klein SL, Flanagan KL. Sex differences in immune responses. Nature Reviews Immunology. 2016 Oct;16(10):626.

25. Giefing-Kröll C, Berger P, Lepperdinger G, Grubeck-Loebenstein B. How sex and age affect immune responses, susceptibility to infections, and response to vaccination. Aging cell. 2015 Jun;14(3):309–21.

26. World Health Organization. Tuberculosis and HIV: some questions and answers. In Tuberculosis and HIV: some questions and answers 1999.

27. Zhao Q, Meng M, Kumar R, Wu Y, Huang J, Lian N, Deng Y, Lin S. The impact of COPD and smoking history on the severity of Covid-19: A systemic review and meta-analysis. Journal of medical virology. 2020 Apr 15.

28. Alraddadi BM, Watson JT, Almarashi A, Abedi GR, Turkistani A, Sadran M, et al. Risk Factors for Primary Middle East Respiratory Syndrome Coronavirus Illness in Humans, Saudi Arabia, 2014. Emerging infectious diseases 2016; 22: 49–55.

29. Badawi A, Ryoo SG. Prevalence of comorbidities in the Middle East respiratory syndrome coronavirus (MERS-CoV): a systematic review and meta-analysis. International Journal of Infectious Diseases. 2016 Aug 1;49:129–33.

